# Home-based Extended Rehabilitation for Older People with Frailty (HERO): a Randomised Controlled Trial

**DOI:** 10.1101/2025.06.17.25329580

**Authors:** Andrew Clegg, Matthew Prescott, Michelle Collinson, Victoria A Goodwin, Ellen Thompson, Rebecca Bestwick, Chris Bojke, David Clarke, Florence Day, Anne Forster, Abi Hall, Claire Hulme, Julie Peacock, Friederike Ziegler, Amanda Farrin

## Abstract

**Objective:** To evaluate whether home-based extended rehabilitation for older people with frailty after hospitalisation with an acute illness or injury can improve physical health-related quality of life and is cost-effective.

**Trial design:** Pragmatic, multi-centre, individually randomised controlled parallel group superiority trial with economic evaluation and embedded process evaluation.

**Setting:** Recruitment from 15 NHS Trusts in England, with home-based intervention delivery.

**Participants:** Eligible participants were 65 years or older with mild/moderate/severe frailty (score of 5-7 on Clinical Frailty Scale) admitted to hospital with acute illness/injury, then discharged home directly, or from intermediate care (post-acute care) rehabilitation services. Recruitment took place December 2017 to August 2021, with follow-up to August 2022.

**Interventions:** Participants were randomly assigned (1.28:1) to the Home-based Older People’s Exercise (HOPE) programme - a 24-week home-based manualised, progressive exercise intervention as extended rehabilitation, or usual care (control). Participants were not masked to allocation.

**Main outcome measures:** Primary outcome was physical health-related quality of life, measured using the physical component summary (PCS) of the modified Short Form 36-item health questionnaire (SF36) at 12 months. Secondary outcomes at six and 12 months included physical and mental health-related quality of life, functional independence, death, hospitalisations and care home admissions. Researchers involved in data collection were masked to allocation.

**Results:** We randomised 740 participants (410 HOPE, 330 control) across 15 sites. 479 (64.7%) participants completed 12-month follow-up. 188 HOPE participants (45.9%) completed 24 weeks of intervention delivery. Over half of participants completed more than 75% of prescribed exercises.

Intention-to-treat analyses showed no evidence that HOPE was superior to control for 12-month PCS score (adjusted mean difference −0.22, 95% CI −1.47 to 1.03; p = 0.73). There was some evidence of a higher rate of all-cause hospitalisations in the control arm (incidence rate ratio 1.12, 95% CI 1.00 to 1.25; p = 0.05), but no differences in other outcomes. The process evaluation found the intervention was largely delivered as intended and proved acceptable to most participants. The economic analysis showed HOPE plus usual care costs of GB£1,401 with 0.024 QALY improvement compared to the control. Incremental cost-effectiveness ratio GB£58,375.

**Limitations:** The HERO trial was delivered during especially challenging circumstances that included the COVID-19 pandemic. We examined outcomes taking account of this but detected no difference in primary or secondary outcomes, providing reassurance that COVID-19 was unlikely to have influenced trial results.

**Conclusions:** Based on our findings, we do not recommend routine commissioning of extended rehabilitation for older people with frailty after discharge home from hospital or intermediate care, following an acute admission with a medical illness or injury.

**Trial registration:** ISRCTN-13927531 (19/04/2017).

## Introduction

Population ageing is accelerating worldwide and maintaining the independence and quality of life of this growing older population is a shared global priority (1, 2). Frailty is an especially problematic expression of population ageing that is characterised by loss of biological reserves and resulting vulnerability to adverse outcomes such as loss of independence, hospitalisation, care home admission and death (3). It is a common condition, with global estimates indicating frailty affects around 12% of community-dwelling people aged 65 years and over, rising to around half of people aged 90 years and over (4). Around 50% of older people in hospital have frailty and are at higher risk of readmission or death following discharge home, compared to robust older people (5, 6). Frailty negatively impacts on quality of life, caregiver burden and health and social care use (7). It accounts for £6billion of annual UK National Health Service (NHS) expenditure (8) and is the strongest predictor of social care costs (9).

In the UK, following acute hospitalisation, around one third of older people with frailty are likely to return home after a brief period of rehabilitation on a hospital ward. A further third are likely to die in hospital or require nursing home care after discharge. The final third are likely to require and receive a longer period of rehabilitation (10). This rehabilitation, termed ‘intermediate care’ (IC), comprises a range of rehabilitation services to promote recovery of independence and reduce the premature need for long-term care – similar to ‘post-acute’ care services internationally. Services typically include provision of rehabilitation in bed-based IC services (e.g. community rehabilitation hospital setting), or via home-based services (e.g. supported discharge teams or community therapy services). UK guidelines recommend a relatively brief contact with these services of two to six weeks (11). The average length of IC stay is around 30 days with 80% of service recipients discharged from IC within six weeks (12). Many recipients of IC do not feel ready to leave the service, indicating possible incomplete recovery. Although IC reduces readmission to hospital within a month of discharge (13), this benefit attenuates, with no reduction in readmissions to hospital over the longer term (14, 15).

A key challenge for healthcare systems internationally is how to sustain the benefit from a short period of rehabilitation in IC over a longer time-horizon. Systematic review evidence indicates exercise interventions based on progressive strength training can improve mobility and function for frail older people and slow progression to disability (16–18). The main objective for this trial was to evaluate the clinical and cost-effectiveness of a programme of progressive strength exercise targeted at key muscle groups for functional mobility, with integrated behaviour change techniques, as extended rehabilitation for older people with frailty after hospitalisation with an acute illness or injury.

## Methods

### Study design and participants

The Home-based Extended Rehabilitation for Older people (HERO) trial was a pragmatic, multi-centre, individually randomised controlled parallel group trial with a two-level partially nested hierarchical design. The trial included an internal pilot and embedded process and economic evaluations.

HERO recruited participants from 15 English NHS hospital trusts within Yorkshire and South West England. The HERO trial protocol has been published (19) and full details of the pre-specified internal pilot criteria, internal pilot results and subsequent trial protocol amendments are available in the supplementary material. Trial amendments were approved by Yorkshire and the Humber (Bradford) Research Ethics Committee (17/YH/0097). The trial was registered with the ISRCTN Registry (13927531) and was conducted in accordance with the principles of Good Clinical Practice and the Declaration of Helsinki.

Eligible patients were 65 years or older admitted to general/elderly medicine or trauma and orthopaedic hospital wards following acute illness or injury, then discharged home directly, or from IC services (bed-based and home based). Patients had mild, moderate or severe frailty, defined as a score of five to seven on the 9-item Clinical Frailty Scale (20) and had to complete the Timed Up and Go Test (TUGT) (21) without additional support. Patients were ineligible if they were permanent care home residents; had significant cognitive impairment (Montreal Cognitive Assessment (MoCA) test <20) (22); had recent (<3 months pre-randomisation) myocardial infarction or unstable angina; had very severe frailty (scored 8 on CFS); were receiving palliative care; had a referral at discharge for condition-specific rehabilitation e.g. pulmonary/stroke rehabilitation or falls prevention programme; had another household member in the trial or were currently participating in another conflicting trial.

Potentially eligible patients were identified via screening on admission to the relevant ward/service by local research staff and through discussions with ward staff. Eligible patients were approached approximately 48h prior to discharge, assessed for capacity to provide informed consent and introduced to the trial verbally. Interested patients were provided with verbal and written information, given appropriate time to consider involvement and where consent was obtained, final eligibility and baseline assessments were completed.

### Randomisation and masking

Participants were randomly assigned in a 1.28:1 ratio to receive the Home-based Older People’s Exercise (HOPE) programme plus usual care, or usual care alone (control) via an online/telephone-based randomisation system at the University of Leeds Clinical Trials Research Unit. Allocation, via a computer-generated minimisation programme, incorporated a random element to ensure arms were balanced for recruiting site, discharge setting (hospital, bed-based IC, home based IC), intended level of HOPE programme (level one, two or three) and reason for admission (acute illness or injury). Due to the nature of HOPE, participants and those involved in HOPE delivery were not masked to treatment allocation. General practitioners, although aware of trial participation, were not informed of allocation. Other health and social care teams remained unaware of both trial participation and allocation. Researchers involved in data collection were masked to allocation with alternative researchers conducting subsequent follow-up assessments if treatment allocation became known. The analysis team were not masked to allocation, however a detailed statistical analysis plan was written and agreed with the research team and independent members of the Trial Steering Committee (TSC) prior to database lock.

### Procedures

The original development of the HOPE programme through co-design has been previously described (20). A detailed description of HOPE delivered during the trial is provided in the Template for Intervention Description and Replication (TIDieR) checklist (supplementary material), with a concise overview provided below.

HOPE, designed specifically for older people with frailty, is a home-based manualised, progressive exercise intervention, graded into three levels of increasing difficulty to account for the spectrum of frailty. The programme aims to improve strength, endurance and balance for basic mobility skills including getting out of bed, standing up from a chair, walking a short distance and getting on/off the toilet. For this trial the original HOPE programme was extended from 12 to 24 weeks to incorporate 12 weeks telephone-based support for intervention sustainability.

HOPE was delivered by trained NHS community physiotherapy teams via scheduled weekly face-to-face or telephone contacts, with the first planned contact within three weeks of discharge home. Participants were provided with a tailored and manualised exercise programme from one of three HOPE programme level manuals, based on their performance on the timed-up-and-go test.

Participants were encouraged to exercise three times daily (approx. 30mins in total) for five days per week as able, with flexibility to allow tailoring to individual needs. Therapists supported participants to appropriately progress their exercise programme through increased frequency, intensity, volume and number/type of difference exercises. The therapists employed behaviour change strategies appropriate to each participant. These included, but were not limited to, promotion of self-monitoring (exercise diary) goal setting, and action planning to meet identified goals, which were reviewed at subsequent contact points.

Usual care was not restricted in either arm and was expected to vary dependent upon participant’s level of health and social care need. Usual care service use was collected for 12-months post-randomisation in both arms.

### Outcomes

Participants completed outcome measures at baseline, six and 12-months via self-report postal questionnaires. Reminder letters were sent to non-responders at three weeks to prompt completion, followed by a telephone call to offer support and/or collect the data over the telephone or to arrange a face-to-face home visit.

The primary outcome was the Physical Component Summary (PCS) score of the Short-Form 36-item Health Questionnaire (SF36) as a measure of physical health-related quality of life (23), measured at 12-months post-randomisation. PCS score was also measured at six months post-randomisation. Other secondary outcomes, measured at six and 12-months post-randomisation, included the SF36 Mental Component Summary (MCS) score, activities of daily living (measured by the Barthel Index of activities of daily living (ADL) (24) and the Nottingham Extended ADL (NEADL) index (25)), health-related quality of life (measured by the EuroQol 5-Dimension 5-Level Version (EQ-5D-5L) (26)) and healthcare resource use. Research staff recorded care home admission status. Hospital readmission rates, all-cause hospitalisation, and hospitalisation due to falls or fractures were recorded using routine Hospital Episodes Statistics (HES) and mortality using linked Office for National Statistics (ONS) data. The primary and secondary outcomes are defined in full in the supplementary material.

Usual care data was collected for 12-months post-randomisation by site research teams from electronic site databases. Adverse events, including death and hospitalisation rates due to falls and/or fracture were collected by sites at five and 11-months post-randomisation. These timepoints were selected to incorporate a status check of participants prior to contact for questionnaires at six and 12 months, reducing the risk of contacting participants who had died.

### Statistical analysis

The sample size of 742 participants (417 HOPE, 325 control) provided 90% power to detect a minimum clinically important difference of three points in the PCS, at the two-sided 5% significance level, allowing for 35% loss to follow-up (27). We assumed a mean PCS score of 30 (SD 9.47) (28), clustering in the HOPE arm only, an average cluster size of seven participants per therapist, an intraclass correlation coefficient of 0.03 (29, 30), and a coefficient of variation in cluster size of 0.7 to account for a varying number of participants per therapist.

All analyses were conducted in SAS version 9.4, for the intention-to-treat population, according to randomisation and regardless of compliance with, or withdrawal from, the trial. A single, final analysis of outcomes (including all internal pilot data) was conducted after data-lock at the end of the trial. With the exception of SF36, participant-reported outcomes were scored according to user-guides, with missing item-level data handled according to guidance where available or imputed using the half-rule (31). SF36 raw scores were scored and handled as per guidance using OPTUM PRO CoRE software (32). All statistical testing was performed at the 5% significance level.

We analysed the primary outcome using a generalised linear mixed-effects regression model, adjusting for the minimisation factors, participant characteristics (age, sex, previous engagement or referral to community rehabilitation services, Charlson index) and PCS score at baseline to test for differences in PCS score between arms at 12-months. We planned to use a partially clustered model to account for clustering of outcomes in the HOPE arm due to therapist effects (33), however this was not possible as the average cluster size was too low. Missing data patterns were explored and multiple imputation via pattern-mixture modelling with 40 imputations was used to impute missing values (including missing data due to death) (34). Treatment allocation, age, sex, ethnicity, discharge setting, reason for admission, intended level of HOPE programme, involvement in rehabilitation programme and baseline PCS score were included in the imputation model. Rubin’s rules were used to combine results of identical analyses performed on each of the imputed datasets (35). Robustness of primary outcome analysis was tested via sensitivity analyses of participants with complete data; excluding participants who had died; and including an additional covariate denoting the period of outcome measure completion (pre-/post-COVID-19 lockdown). Results were expressed as adjusted mean differences with 95% CIs and p values.

Continuous secondary outcomes at months six and 12 were analysed as per the primary outcome and adjusted for minimisation factors, participant characteristics and respective baseline score. Sensitivity analyses of participants with complete data were compared with analyses using multiple imputations for these outcomes. Binary secondary outcomes at 12-months were analysed similarly in logistic (care home admission, hospital readmission, mortality) or Poisson (all-cause hospitalisations, hospitalisations due to falls) regression models, using available data without multiple imputation, expressing results as odds ratios (ORs) or rate ratios (RR), where relevant, with 95% CIs. Time to death was analysed using Cox proportional-hazards, adjusting for the minimisation factors and participant characteristics. Assumptions were checked for all regression models using residual plots (linear, logistic and Poisson) or via assessment of proportional hazards (Cox).

Intervention delivery was summarised descriptively. A complier average causal effect (CACE) analysis (36) was undertaken to understand the impact of participant and therapist compliance on the primary outcome using a two-stage instrumental variable regression approach with randomised arm as the instrumental variable, adjusting for baseline PCS score, age, gender and level of previous engagement with community rehabilitation services. Compliance was defined in four ways using combinations of the number of home visits and percentage completion of all exercises prescribed (full details in supplementary material).

The number of participants withdrawing from trial elements was summarised by arm with reasons, where available. Safety data relating to deaths and hospitalisations resulting from falls and/or fractures were summarised descriptively by arm. The TSC and Trial Management Group (TMG) reviewed accumulating safety data at agreed intervals throughout the trial.

### Health economic analysis

The within trial cost-effectiveness analyses, guided by the recommendations of the National Institute for Health and Care Excellence (37), compared HOPE to control. The analyses took the perspective of the health and social care sectors using quality adjusted life years (QALYs) derived from the EQ-5D-5L and survival data. Costs included participant-utilised resources and were valued using national prices sources. Participants were followed for 12 months. The analyses used incremental cost-effectiveness ratios (ICERs) and were conducted with and without multiple imputation (MI). A Decision-Analytical Model (DAM) was also constructed to extrapolate results over the longer-term (up to 15 years). Transition probabilities for the DAM were estimated using parametric survival and regression analyses of the trial data. Results from the model are also presented as ICERs.

### Process evaluation

Qualitative mixed-methods process evaluation incorporating non-participant observations of intervention delivery; semi-structured interviews of intervention and usual care participants, therapy services managers, physiotherapists and therapy assistants; and documentary analysis of the therapy records and participant exercise diaries. Fidelity in intervention delivery and acceptability of its receipt and delivery were explored. Data analysis was based on thematic and underpinned by Normalisation Process Theory (38).

### Patient and public involvement

Patient and public contributors were involved during the co-design of the original HOPE programme, and throughout the trial, providing invaluable contributions to design, documentation and outputs. PPI representatives were involved in trial management and governance through representation on the Trial Management Group and independent Trial Steering Committee.

## Results

Between December 01, 2017, and August 09, 2021, 16687 patients were screened, 5505 (33.0%) were deemed eligible for approach, of which 905 (16.4%) consented to further eligibility checks and 775 (85.6% of consented) were eligible (Figure 1). We randomised 740 (95.5% of eligible) participants to receive HOPE (410) or control (330). Randomised participants were similar in age to those screened, however some differences were noted for gender, ethnicity and reason for hospital admission (supplementary material). Recruitment paused during the COVID-19 pandemic between March 16, to October 29, 2020, in accordance with national guidance, and restarted on October 30, 2020. Follow-up ended on September 01, 2022, with 539 (72.8%%) participants completing six-month follow-up (299 [72.9%] of 410 in HOPE, 240 [72.7%] of 330 in control) and 479 (64.7%) participants completing 12-month follow-up (264 [64.4%] of 410 in HOPE, 215 [65.2%] of 330 in control (Figure 1). Summaries of eligibility violations and withdrawals are provided in the supplementary material. All randomised participants were included in the intention-to-treat analysis.

**Figure 1.**
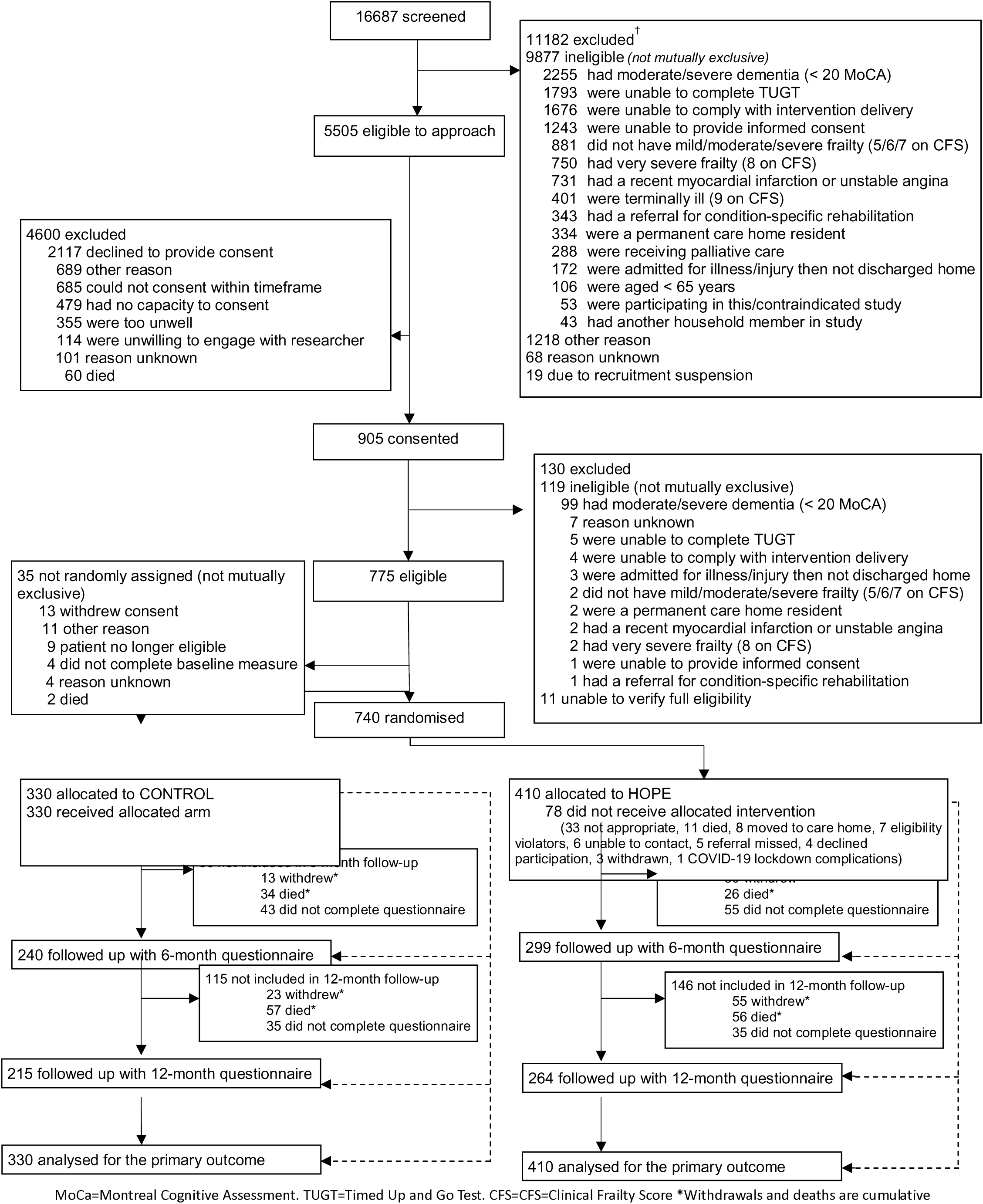
Trial profile.

Baseline demographics and characteristics were broadly similar between the arms (Table 1). In the 740 randomised participants with available data, the mean age was 82.6 years (SD 7.1 years), 486 (65.7%) were female, and 699 (97.6%) were white. Over two-thirds of participants (511 [69.1%]) were admitted to hospital due to acute illness and 300 (40.5%) were discharged from home-based intermediate care. The majority of participants had either mild frailty (375 [50.8%]) or moderate frailty (324 [43.9%]) and were allocated to HOPE Level 1 (466 [63.0%]). Participant reported measures were balanced between the arms.

**Table 1:**
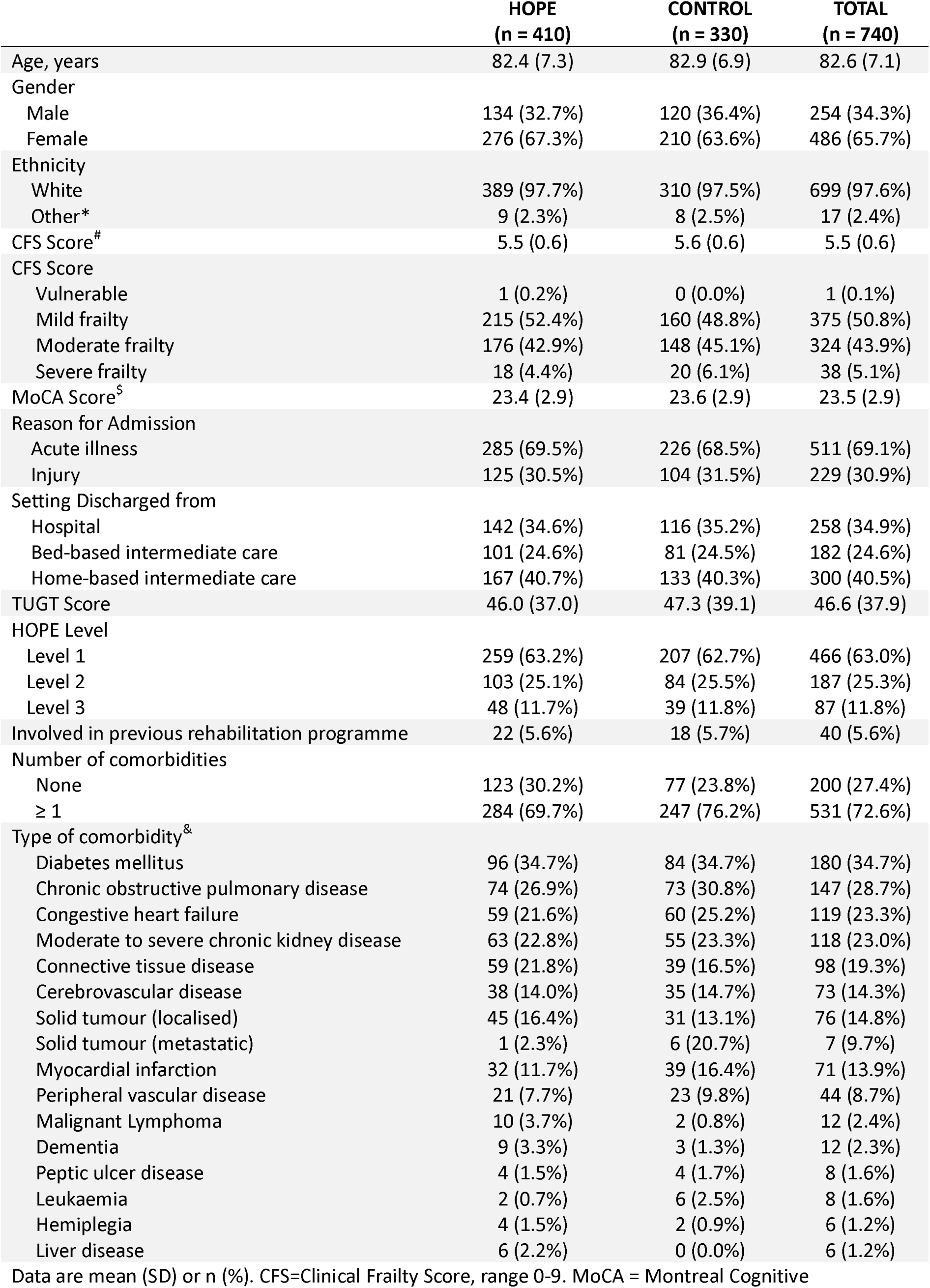
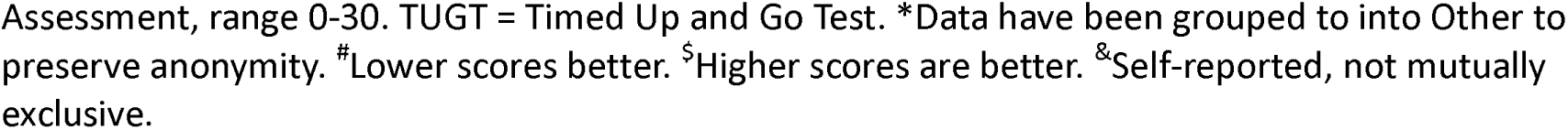
Baseline characteristics.

|  | **HOPE**<br>**(n = 410)** | **CONTROL**<br>**(n = 330)** | **TOTAL**<br>**(n = 740)** |
| --- | --- | --- | --- |
| Age, years | 82.4 (7.3) | 82.9 (6.9) | 82.6 (7.1) |
| Gender |  |  |  |
| Male | 134 (32.7%) | 120 (36.4%) | 254 (34.3%) |
| Female | 276 (67.3%) | 210 (63.6%) | 486 (65.7%) |
| Ethnicity |  |  |  |
| White | 389 (97.7%) | 310 (97.5%) | 699 (97.6%) |
| Other* | 9 (2.3%) | 8 (2.5%) | 17 (2.4%) |
| CFS Score<sup>#</sup> | 5.5 (0.6) | 5.6 (0.6) | 5.5 (0.6) |
| CFS Score |  |  |  |
| Vulnerable | 1 (0.2%) | 0 (0.0%) | 1 (0.1%) |
| Mild frailty | 215 (52.4%) | 160 (48.8%) | 375 (50.8%) |
| Moderate frailty | 176 (42.9%) | 148 (45.1%) | 324 (43.9%) |
| Severe frailty | 18 (4.4%) | 20 (6.1%) | 38 (5.1%) |
| MoCA Score<sup>§</sup> | 23.4 (2.9) | 23.6 (2.9) | 23.5 (2.9) |
| Reason for Admission |  |  |  |
| Acute illness | 285 (69.5%) | 226 (68.5%) | 511 (69.1%) |
| Injury | 125 (30.5%) | 104 (31.5%) | 229 (30.9%) |
| Setting Discharged from |  |  |  |
| Hospital | 142 (34.6%) | 116 (35.2%) | 258 (34.9%) |
| Bed-based intermediate care | 101 (24.6%) | 81 (24.5%) | 182 (24.6%) |
| Home-based intermediate care | 167 (40.7%) | 133 (40.3%) | 300 (40.5%) |
| TUGT Score | 46.0 (37.0) | 47.3 (39.1) | 46.6 (37.9) |
| HOPE Level |  |  |  |
| Level 1 | 259 (63.2%) | 207 (62.7%) | 466 (63.0%) |
| Level 2 | 103 (25.1%) | 84 (25.5%) | 187 (25.3%) |
| Level 3 | 48 (11.7%) | 39 (11.8%) | 87 (11.8%) |
| Involved in previous rehabilitation programme | 22 (5.6%) | 18 (5.7%) | 40 (5.6%) |
| Number of comorbidities |  |  |  |
| None | 123 (30.2%) | 77 (23.8%) | 200 (27.4%) |
| ≥ 1 | 284 (69.7%) | 247 (76.2%) | 531 (72.6%) |
| Type of comorbidity<sup>&</sup> |  |  |  |
| Diabetes mellitus | 96 (34.7%) | 84 (34.7%) | 180 (34.7%) |
| Chronic obstructive pulmonary disease | 74 (26.9%) | 73 (30.8%) | 147 (28.7%) |
| Congestive heart failure | 59 (21.6%) | 60 (25.2%) | 119 (23.3%) |
| Moderate to severe chronic kidney disease | 63 (22.8%) | 55 (23.3%) | 118 (23.0%) |
| Connective tissue disease | 59 (21.8%) | 39 (16.5%) | 98 (19.3%) |
| Cerebrovascular disease | 38 (14.0%) | 35 (14.7%) | 73 (14.3%) |
| Solid tumour (localised) | 45 (16.4%) | 31 (13.1%) | 76 (14.8%) |
| Solid tumour (metastatic) | 1 (2.3%) | 6 (20.7%) | 7 (9.7%) |
| Myocardial infarction | 32 (11.7%) | 39 (16.4%) | 71 (13.9%) |
| Peripheral vascular disease | 21 (7.7%) | 23 (9.8%) | 44 (8.7%) |
| Malignant Lymphoma | 10 (3.7%) | 2 (0.8%) | 12 (2.4%) |
| Dementia | 9 (3.3%) | 3 (1.3%) | 12 (2.3%) |
| Peptic ulcer disease | 4 (1.5%) | 4 (1.7%) | 8 (1.6%) |
| Leukaemia | 2 (0.7%) | 6 (2.5%) | 8 (1.6%) |
| Hemiplegia | 4 (1.5%) | 2 (0.9%) | 6 (1.2%) |
| Liver disease | 6 (2.2%) | 0 (0.0%) | 6 (1.2%) |

Data are mean (SD) or n (%). CFS=Clinical Frailty Score, range 0-9. MoCA = Montreal Cognitive
Assessment, range 0-30. TUGT = Timed Up and Go Test. \*Data have been grouped to into Other to preserve anonymity. #Lower scores better. §Higher scores are better. &Self-reported, not mutually exclusive.

Of the 410 participants allocated to HOPE, 332 (81.0%) started the intervention and had an initial home visit, 223 (54.4%) had at least five home visits and 188 (45.9%) completed 24 weeks of intervention delivery (supplementary material). Over half of participants completed more than 75% of prescribed exercises.

Unadjusted and adjusted mean scores for the primary outcome, PCS score, were stable over time and similar between arms at both six and 12-months (Table 2 and 3, Figure 2). There was no evidence that HOPE was superior to control for PCS score at 12-months (primary endpoint, adjusted mean difference −0.22, 95% CI −1.47 to 1.03; p = 0.73) or at six months (adjusted mean difference −1.10, 95% CI −2.32 to 0.12; p=0.08, Table 3). All sensitivity analyses provided conclusions consistent with the primary analysis.

**Figure 2.**
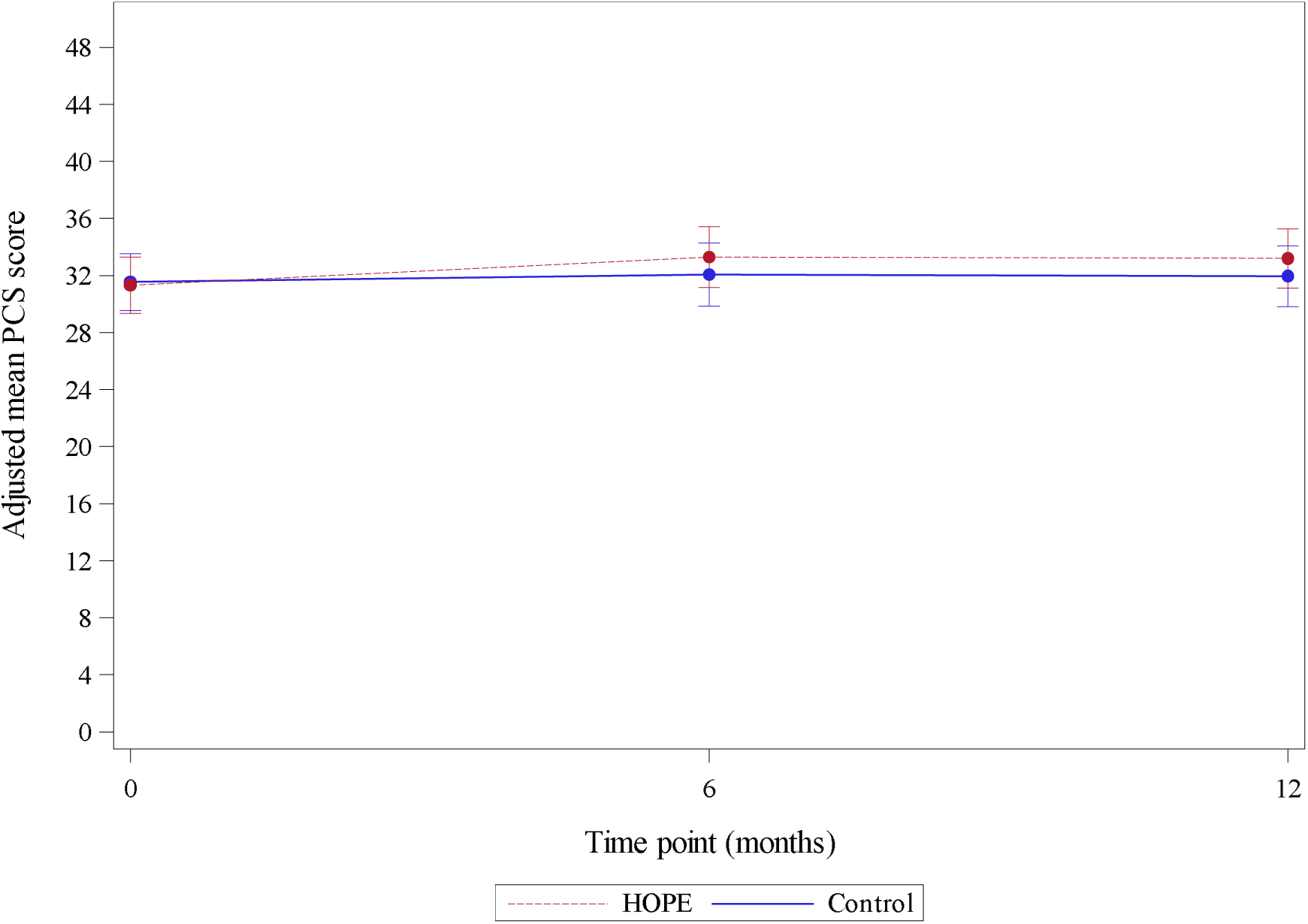
Health-related quality of life during follow-up, measured by adjusted SF36 PCS. SF36 PCS= SF36 Physical Component Score, range 0-100. Estimated from a linear regression, adjusted for the stratification factors, age, previous engagement or referral to community rehabilitation services, charlson comorbidity index and baseline PCS score. Error bars depict 95% CIs.

**Table 2.**
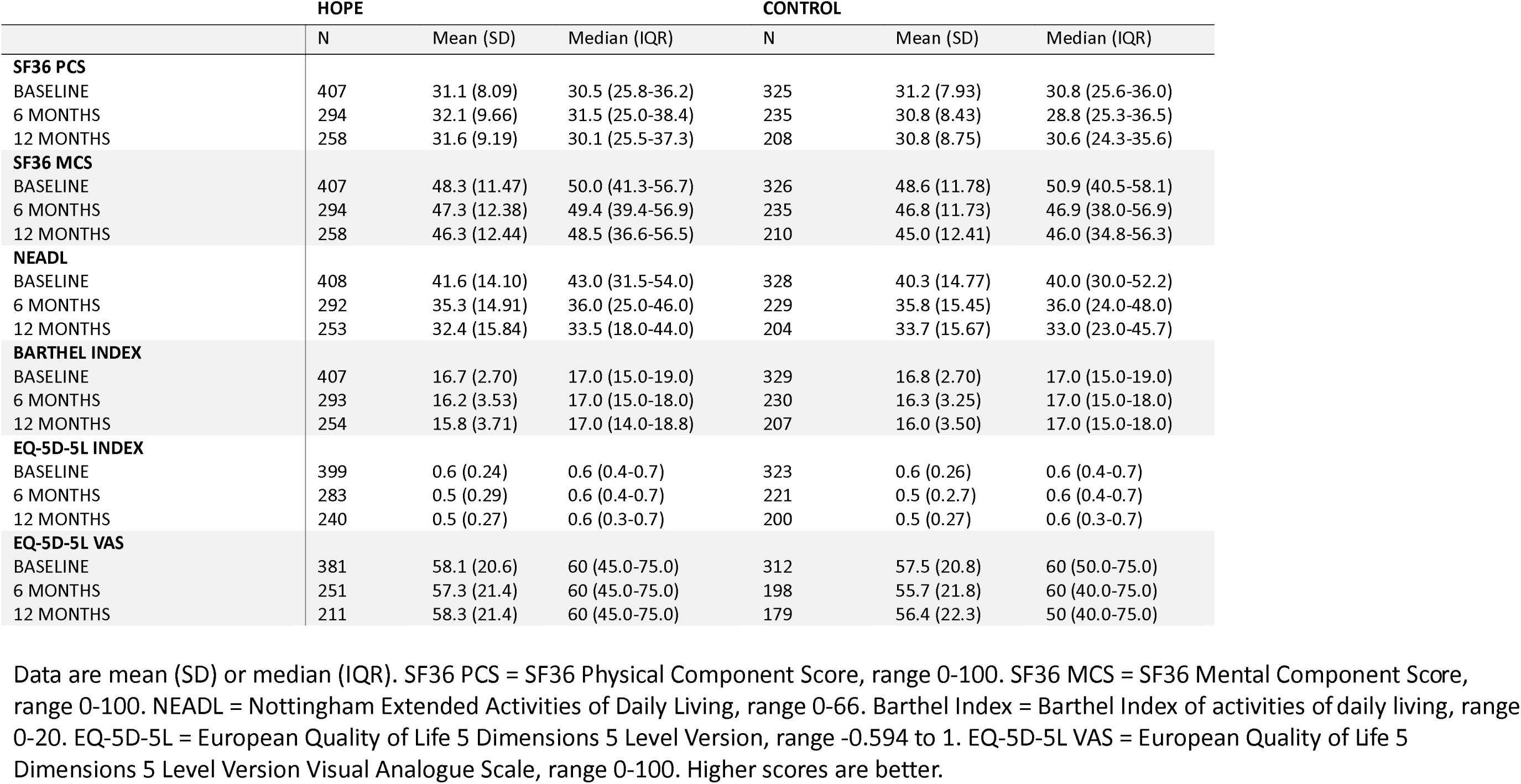
Raw summaries of patient-reported outcome measures by trial arm and timepoint.

|  | HOPE |  |  | CONTROL |  |  |
| --- | --- | --- | --- | --- | --- | --- |
|  | N | Mean (SD) | Median (IQR) | N | Mean (SD) | Median (IQR) |
| **SF36 PCS** |  |  |  |  |  |  |
| BASELINE | 407 | 31.1 (8.09) | 30.5 (25.8-36.2) | 325 | 31.2 (7.93) | 30.8 (25.6-36.0) |
| 6 MONTHS | 294 | 32.1 (9.66) | 31.5 (25.0-38.4) | 235 | 30.8 (8.43) | 28.8 (25.3-36.5) |
| 12 MONTHS | 258 | 31.6 (9.19) | 30.1 (25.5-37.3) | 208 | 30.8 (8.75) | 30.6 (24.3-35.6) |
| **SF36 MCS** |  |  |  |  |  |  |
| BASELINE | 407 | 48.3 (11.47) | 50.0 (41.3-56.7) | 326 | 48.6 (11.78) | 50.9 (40.5-58.1) |
| 6 MONTHS | 294 | 47.3 (12.38) | 49.4 (39.4-56.9) | 235 | 46.8 (11.73) | 46.9 (38.0-56.9) |
| 12 MONTHS | 258 | 46.3 (12.44) | 48.5 (36.6-56.5) | 210 | 45.0 (12.41) | 46.0 (34.8-56.3) |
| **NEADL** |  |  |  |  |  |  |
| BASELINE | 408 | 41.6 (14.10) | 43.0 (31.5-54.0) | 328 | 40.3 (14.77) | 40.0 (30.0-52.2) |
| 6 MONTHS | 292 | 35.3 (14.91) | 36.0 (25.0-46.0) | 229 | 35.8 (15.45) | 36.0 (24.0-48.0) |
| 12 MONTHS | 253 | 32.4 (15.84) | 33.5 (18.0-44.0) | 204 | 33.7 (15.67) | 33.0 (23.0-45.7) |
| **BARTHEL INDEX** |  |  |  |  |  |  |
| BASELINE | 407 | 16.7 (2.70) | 17.0 (15.0-19.0) | 329 | 16.8 (2.70) | 17.0 (15.0-19.0) |
| 6 MONTHS | 293 | 16.2 (3.53) | 17.0 (15.0-18.0) | 230 | 16.3 (3.25) | 17.0 (15.0-18.0) |
| 12 MONTHS | 254 | 15.8 (3.71) | 17.0 (14.0-18.8) | 207 | 16.0 (3.50) | 17.0 (15.0-18.0) |
| **EQ-5D-5L INDEX** |  |  |  |  |  |  |
| BASELINE | 399 | 0.6 (0.24) | 0.6 (0.4-0.7) | 323 | 0.6 (0.26) | 0.6 (0.4-0.7) |
| 6 MONTHS | 283 | 0.5 (0.29) | 0.6 (0.4-0.7) | 221 | 0.5 (0.2.7) | 0.6 (0.4-0.7) |
| 12 MONTHS | 240 | 0.5 (0.27) | 0.6 (0.3-0.7) | 200 | 0.5 (0.27) | 0.6 (0.3-0.7) |
| **EQ-5D-5L VAS** |  |  |  |  |  |  |
| BASELINE | 381 | 58.1 (20.6) | 60 (45.0-75.0) | 312 | 57.5 (20.8) | 60 (50.0-75.0) |
| 6 MONTHS | 251 | 57.3 (21.4) | 60 (45.0-75.0) | 198 | 55.7 (21.8) | 60 (40.0-75.0) |
| 12 MONTHS | 211 | 58.3 (21.4) | 60 (45.0-75.0) | 179 | 56.4 (22.3) | 50 (40.0-75.0) |

Data are mean (SD) or median (IQR). SF36 PCS = SF36 Physical Component Score, range 0-100. SF36 MCS = SF36 Mental Component Score, range 0-100. NEADL = Nottingham Extended Activities of Daily Living, range 0-66. Barthel Index = Barthel Index of activities of daily living, range 0-20. EQ-5D-5L = European Quality of Life 5 Dimensions 5 Level Version, range -0.594 to 1. EQ-5D-5L VAS = European Quality of Life 5 Dimensions 5 Level Version Visual Analogue Scale, range 0-100. Higher scores are better.

**Table 3.**
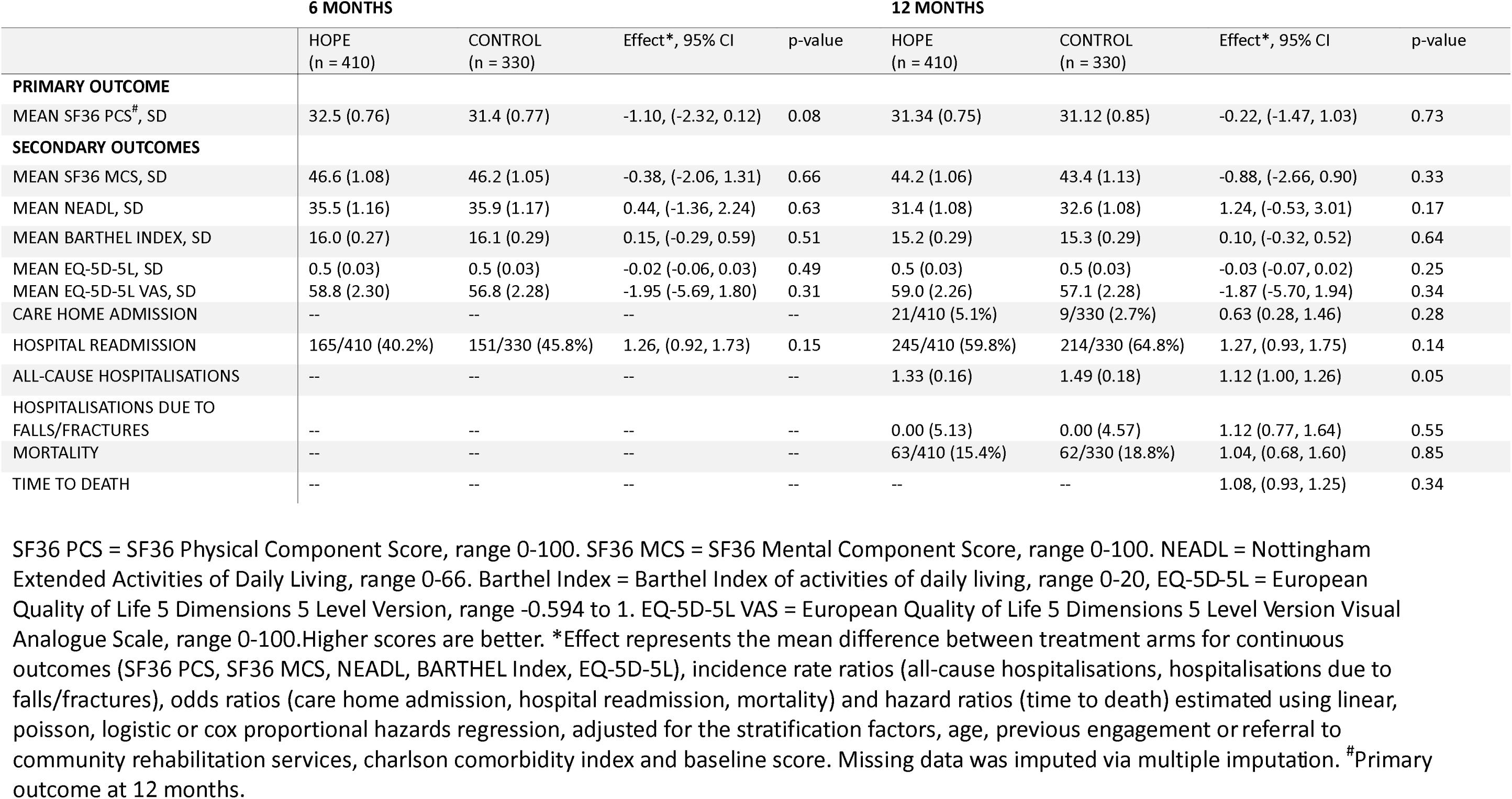
Primary outcome at 12 months and secondary outcomes at 6 and 12 months.

|  | 6 MONTHS |  |  |  | 12 MONTHS |  |  |  |
| --- | --- | --- | --- | --- | --- | --- | --- | --- |
|  | HOPE<br>(n = 410) | CONTROL<br>(n = 330) | Effect*, 95% CI | p-value | HOPE<br>(n = 410) | CONTROL<br>(n = 330) | Effect*, 95% CI | p-value |
| **PRIMARY OUTCOME** |  |  |  |  |  |  |  |  |
| MEAN SF36 PCS<sup>#</sup>, SD | 32.5 (0.76) | 31.4 (0.77) | -1.10, (-2.32, 0.12) | 0.08 | 31.34 (0.75) | 31.12 (0.85) | -0.22, (-1.47, 1.03) | 0.73 |
| **SECONDARY OUTCOMES** |  |  |  |  |  |  |  |  |
| MEAN SF36 MCS, SD | 46.6 (1.08) | 46.2 (1.05) | -0.38, (-2.06, 1.31) | 0.66 | 44.2 (1.06) | 43.4 (1.13) | -0.88, (-2.66, 0.90) | 0.33 |
| MEAN NEADL, SD | 35.5 (1.16) | 35.9 (1.17) | 0.44, (-1.36, 2.24) | 0.63 | 31.4 (1.08) | 32.6 (1.08) | 1.24, (-0.53, 3.01) | 0.17 |
| MEAN BARTHEL INDEX, SD | 16.0 (0.27) | 16.1 (0.29) | 0.15, (-0.29, 0.59) | 0.51 | 15.2 (0.29) | 15.3 (0.29) | 0.10, (-0.32, 0.52) | 0.64 |
| MEAN EQ-5D-5L, SD | 0.5 (0.03) | 0.5 (0.03) | -0.02 (-0.06, 0.03) | 0.49 | 0.5 (0.03) | 0.5 (0.03) | -0.03 (-0.07, 0.02) | 0.25 |
| MEAN EQ-5D-5L VAS, SD | 58.8 (2.30) | 56.8 (2.28) | -1.95 (-5.69, 1.80) | 0.31 | 59.0 (2.26) | 57.1 (2.28) | -1.87 (-5.70, 1.94) | 0.34 |
| CARE HOME ADMISSION | -- | -- | -- | -- | 21/410 (5.1%) | 9/330 (2.7%) | 0.63 (0.28, 1.46) | 0.28 |
| HOSPITAL READMISSION | 165/410 (40.2%) | 151/330 (45.8%) | 1.26, (0.92, 1.73) | 0.15 | 245/410 (59.8%) | 214/330 (64.8%) | 1.27, (0.93, 1.75) | 0.14 |
| ALL-CAUSE HOSPITALISATIONS | -- | -- | -- | -- | 1.33 (0.16) | 1.49 (0.18) | 1.12 (1.00, 1.26) | 0.05 |
| HOSPITALISATIONS DUE TO FALLS/FRACTURES | -- | -- | -- | -- | 0.00 (5.13) | 0.00 (4.57) | 1.12 (0.77, 1.64) | 0.55 |
| MORTALITY | -- | -- | -- | -- | 63/410 (15.4%) | 62/330 (18.8%) | 1.04, (0.68, 1.60) | 0.85 |
| TIME TO DEATH | -- | -- | -- | -- | -- | -- | 1.08, (0.93, 1.25) | 0.34 |

SF36 PCS = SF36 Physical Component Score, range 0-100. SF36 MCS = SF36 Mental Component Score, range 0-100. NEADL = Nottingham Extended Activities of Daily Living, range 0-66. Barthel Index = Barthel Index of activities of daily living, range 0-20, EQ-5D-5L = European Quality of Life 5 Dimensions 5 Level Version, range -0.594 to 1. EQ-5D-5L VAS = European Quality of Life 5 Dimensions 5 Level Version Visual Analogue Scale, range 0-100. Higher scores are better. \*Effect represents the mean difference between treatment arms for continuous outcomes (SF36 PCS, SF36 MCS, NEADL, BARTHEL Index, EQ-5D-5L), incidence rate ratios (all-cause hospitalisations, hospitalisations due to falls/fractures), odds ratios (care home admission, hospital readmission, mortality) and hazard ratios (time to death) estimated using linear, poisson, logistic or cox proportional hazards regression, adjusted for the stratification factors, age, previous engagement or referral to community rehabilitation services, charlson comorbidity index and baseline score. Missing data was imputed via multiple imputation. <sup>#</sup>Primary outcome at 12 months.

On average, unadjusted and adjusted scores for MCS (mental health, vitality and social functioning), NEADL and Barthel Index (activities of daily living) and EQ-5D-5L (health-related quality of life) decreased over time in both arms with the largest reductions noted for the NEADL; there was no evidence of a significant difference in these scores between arms at six or 12-months (Table 2 and 3).

There was no evidence of a significant difference in care home admissions, hospital readmission rates, hospitalisations due to falls, mortality or time to death at 12-months (Table 2). There was some evidence that the rate of all-cause hospitalisations was higher in the control arm than in the HOPE arm (incidence rate ratio 1.12, 95% CI 1.00 to 1.25; p = 0.05) (Table 2). The proportion of deaths was similar across both arms (supplementary material). No related and unexpected serious adverse events were reported.

The CACE analysis found no difference in PCS scores at 12-months when considering varying adherence to home visits and exercises (supplementary material).

### Health economic analysis

Full details of the health economic analysis will be reported separately. At 12 months the mean estimated QALYs and costs were higher for the HOPE arm compared to control (incremental difference 0.024 QALYs and GB£1,401 costs). The ICER was GB£58,375. Similarly, the MI analysis gave an ICER of GB£49,711 (incremental difference 0.03 QALYs and GB£1,469 costs). Both ICERs are above the NICE recommended threshold (37). The results from the longer-term decision-analytical model, applying a five, 10 and 15-year time-horizon, also gave respective ICERs higher than the UK threshold. Thus, HOPE was not shown to be cost-effective.

### Process evaluation

The process evaluation was successfully completed and will be reported in full separately. Key findings were that HOPE was broadly delivered as planned with no significant variation between site/regions. Therapists and intervention participants perceived HOPE to be an acceptable rehabilitation intervention for use in this post-acute context. Some therapists felt that greater clarity about how the intervention could be best be adapted to individual circumstances was needed. With appropriate resource allocation, it was feasible for HOPE to be delivered by therapists and appropriately trained and supervised therapy assistants in community-based rehabilitation settings.

## Discussion

Our trial findings indicate no benefit of extended rehabilitation (via the HOPE programme) for older people with frailty on physical health, mental health, or activities of daily living. There was some evidence for a reduction in all-cause hospitalisations, but no effect on care home admissions or mortality. Overall, when considered collectively, the intervention was not cost-effective.

The HERO trial was delivered during especially challenging circumstances that included the COVID-19 pandemic. We paused trial recruitment at a very early stage of the pandemic, at which point the majority of participants had received the required number of face-to-face intervention visits. However, we remained mindful of the potential impact of COVID-19 restrictions on the physical activity and physical role components of the SF36, which include engagement in outdoor activities. We examined outcomes taking account of this but detected no difference in primary or secondary outcomes, providing reassurance that COVID-19 was unlikely to have influenced trial results.

Our findings indicate that overall, reported intervention adherence was reasonable, taking account of the frailty of the population and day-to-day health fluctuations, with over half of participants completing more than 75% of prescribed exercises. Reliable data on intervention adherence is challenging to collect, and it is possible that overall intervention adherence may have been lower than reported, thus reducing the potential for benefit.

Our HOPE intervention was designed to be tailored to individual needs, allowing flexibility in initial intervention intensity to engage participants who might have struggled with a higher intensity. Although intervention training was developed to account for this, our process evaluation findings included feedback from some therapists that greater flexibility in intervention intensity was required, suggesting a need for a stronger emphasis on this in initial training to support overall fidelity. Our CACE analysis, however, did not show any clear relationship between adherence and intervention effect, indicating that whatever the intervention adherence, it is challenging to generate intervention effect for this particular population. Related to this, some previous exercise interventions for older people have included more challenging components such as using progressive weights or resistance bands, and it is possible that the resistance exercise training in the HOPE programme was not intensive enough to generate intervention effect.

Findings conflict with the robust evidence base for resistance exercise training to improve outcomes for older people with frailty. This evidence has typically been generated from the more stable population of community-dwelling older people outside of the unstable, unpredictable fluctuations that often accompany acute illness or after an injury. It is plausible that the adverse health trajectories of older people with frailty after acute illness or injury, and the accompanying challenges including day to day health fluctuations, general fatigue and weakness, mean that it is more difficult to generate similar benefits in this group when compared with a more general population of community-dwelling older people.

Considered collectively, our trial findings provide important evidence for policymakers and commissioners of rehabilitation services for older people globally. Based on our findings, we do not recommend routine commissioning of extended rehabilitation for older people with frailty after discharge home from hospital or IC, following an acute admission with a medical illness or injury. Instead, we recommend that available resources should be directed towards evidence-based core IC services to meet the needs of the growing population of older people in the UK and internationally, alongside resources for resistance exercise training targeted at the more stable population of community-dwelling older people with frailty.

## Supporting information

Consort Checklist

## Data Availability

Data supporting this work are available on reasonable request. All requests will be reviewed by relevant stakeholders, based on the principles of a controlled access approach. All data requests would be subject to review by a subgroup of the trial team, which will include the chief investigator (AC) and data guarantor (AJF). Access to anonymised data could be granted following this review. All data-sharing activities would require a data-sharing agreement. Requests to access data should be made to the corresponding author (Andrew Clegg)in the first instance.

## Research in context

### Evidence before this study

The HERO trial was funded by the National Institute for Health and Care Research through a commissioned research call following a detailed evidence synthesis that identified an evidence gap relating to extended rehabilitation for older people with frailty. We supplemented this with a further review of the literature searching Medline and Cochrane databases from 2000 (when reference-standard frailty measures were first reported) to March 2024 to evaluate evidence published after the trial commenced. Randomised trials evaluating extended rehabilitation for older adults with frailty discharged home from hospital or linked intermediate care (post-acute care) after admission with acute illness or injury, compared with usual care or no intervention, were considered eligible. All health outcomes were considered. Search terms used four key concepts: frailty, older adults, hospital discharge, extended rehabilitation. Reference lists from identified studies were used to identify further potentially relevant studies. We identified four individual trials and one systematic review exploring extended rehabilitation (either home-based or centre-based), following discharge from acute hospitalisation and standard rehabilitation pathways, in older adult populations. These trials and systematic review were for a general older adult population acutely admitted to hospital with a range of medical issues. These studies were all inclusive of a mixed older adult population, and while these studies will most likely have included individuals with frailty, the populations also included more robust individuals, and none presented data with specific reference to frailty. It is also plausible that drop out from these trial/interventions was not at random and included frailer participants. Although the trials reported positive effects on outcomes including mobility, strength and readmission to hospital, one trial reported an increase in falls in the intervention group. It is difficult to draw conclusions as to the effectiveness of those extended rehabilitation interventions for older adults with frailty.

### Added value of this study

This is the first RCT to evaluate extended rehabilitation following discharge from acute hospitalisation in a population of older adults with well-defined frailty. In contrast to other studies where frailty was not an inclusion criteria, and where the study population was more mixed, the HERO trial did not demonstrate effectiveness of extended rehabilitation for older people with frailty.

### Implications of all the available evidence

Our main findings conflict with the available evidence for home-based extended rehabilitation in mixed older adult populations. The unstable health profile associated with frailty following acute hospitalisation may mean it is more difficult to generate similar benefits in this population. Although the reduction in hospitalisations reported in our trial aligned with findings in other trials, our overall health economic evaluation indicated that the intervention was not cost-effective. Our findings therefore do not support routine commissioning of home exercise-based extended rehabilitation for older people with frailty after they have been discharged from standard rehabilitation in hospital, bed-based intermediate care or home-based intermediate care services.

## Contributors

AC co-conceived and designed the HERO trial and had overall responsibility in his role as Chief Investigator. AJF co-conceived and designed the HERO trial, was responsible for its overall implementation across Leeds Clinical Trials Research Unit, and supervised the statistical analysis. MC provided statistical input into the statistical analysis plan, under the supervision of AJF. ET provided statistical input into the implementation and statistical analysis plan, under the supervision of MC and AJF. VG, AF contributed to the design and implementation of the trial. FD coordinated operational delivery of the trial in the CTRU. MP coordinated trial site setup including intervention training, and site liaison throughout the trial. CH and CB designed the health economic evaluation. RB implemented the health economic analysis plan under the supervision of CB and CH. DJC designed the process evaluation. FZ and AH implemented the process evaluation and analysis under the supervision of DJC and AF. JP provided patient and public advice to inform trial implementation and reporting. AC, MP, MC, VG, ET, RB, CB, DJC, CH, AJF drafted the manuscript. All authors commented on drafts of the paper. All authors have approved the final draft of the manuscript. MC and ET had full access to, and verified, all the data in the study. AC and AJF had final responsibility for the decision to submit for publication. AJF is data guarantor.

## Transparency declaration

The lead author (the manuscript’s guarantor) affirms that the manuscript is an honest, accurate, and transparent account of the study being reported; that no important aspects of the study have been omitted; and that any discrepancies from the study as planned have been explained.

## Data sharing

Data supporting this work are available on reasonable request. All requests will be reviewed by relevant stakeholders, based on the principles of a controlled access approach. All data requests would be subject to review by a subgroup of the trial team, which will include the chief investigator (AC) and data guarantor (AJF). Access to anonymised data could be granted following this review. All data-sharing activities would require a data-sharing agreement. Requests to access data should be made to the corresponding author in the first instance.

## Acknowledgements

The study was funded by the NIHR Health Technology Assessment Programme (15/47/07). The study sponsor was Bradford Teaching Hospitals NHS Foundation Trust. The funder and the sponsor had no role in data collection, analysis, interpretation, writing of the manuscript or the decision to submit for publication.

AC is part-funded by the National Institute for Health and Care Research (NIHR) Applied Research Collaboration Yorkshire & Humber, the NIHR Leeds Biomedical Research Centre, and Health Data Research UK, an initiative funded by UK Research and Innovation Councils, NIHR and the UK devolved administrations and leading medical research charities. VG is funded by the NIHR Applied Research Collaboration South West Peninsula (PenARC). CH is part-funded by PenARC. The views expressed in this publication are those of the authors and not necessarily those of the National Health Service, the NIHR, or the Department of Health and Social Care.

We would like to thank the 740 participants and 15 sites involved in the trial, Rifat Parveen from Carer’s Resource Bradford District, as well as the Trial Steering Committee for their support throughout. This work uses data provided by patients and collected by the NHS as part of their care and support.

We are grateful to former Trial Management Group members Bonnie Cundill, Suzanne Hartley, Amanda Lilley-Kelly, Silviya Nikolova, Catriona Parker, Phil Wright and John Young. We would also like to thank Sian Drake, Alison Ellwood, Dax Everitt, Marie Fletcher, Madeline Goodwin, Sarah Hallas, Michael Holland, Judith Horrocks, Adam Hussain, Ruth Illingworth, Ikhlaq Jacob, Farhat Mahmood, Lubena Mirza, Ismail Patel, Peter Saunders, Claire Simpson, Thomas Smith, Gill Thornton, Jonathon Turner and Ian Wheeler as additional research staff who worked on the HERO trial.

## License statement

The Corresponding Author has the right to grant on behalf of all authors and does grant on behalf of all authors, an exclusive licence (or non exclusive for government employees) on a worldwide basis to the BMJ Publishing Group Ltd to permit this article (if accepted) to be published in BMJ editions and any other BMJPGL products and sublicences such use and exploit all subsidiary rights, as set out in our licence.

## References

1. Decade of Healthy Ageing 2021-2030: Plan of Action. United Nations, 2020.

2. Whitty C. Chief Medical Officer’s Annual Report 2023: Health in an Ageing Society. 2023.

3. Clegg A, Young J, Iliffe S, Rikkert MO, Rockwood K. Frailty in elderly people. Lancet. 2013;381(9868):752–62.

4. O’Caoimh R, Sezgin D, O’Donovan MR, Molloy DW, Clegg A, Rockwood K, Liew A. Prevalence of frailty in 62 countries across the world: a systematic review and meta-analysis of population-level studies. Age Ageing. 2021;50(1):96–104.

5. Kahlon S, Pederson J, Majumdar SR, Belga S, Lau D, Fradette M, Boyko D, Bakal JA, Johnston C, Padwal RS, McAlister FA. Association between frailty and 30-day outcomes after discharge from hospital. CMAJ. 2015;187(11):799–804.

6. Keeble E, Roberts HC, Williams CD, Van Oppen J, Conroy SP. Outcomes of hospital admissions among frail older people: a 2-year cohort study. Br J Gen Pract. 2019;69(685):e555–e60.

7. Clegg A, Young J, Iliffe S, Rikkert MO, Rockwood K. Frailty in elderly people. The lancet. 2013;381(9868):752–62.

8. Han L, Clegg A, Doran T, Fraser L. The impact of frailty on healthcare resource use: a longitudinal analysis using the Clinical Practice Research Datalink in England. Age Ageing. 2019;48(5):665–71.

9. Nikolova S, Heaven A, Hulme C, West R, Pendleton N, Humphrey S, Cundill B, Clegg A. Social care costs for community-dwelling older people living with frailty. Health and Social Care in the Community. 2022;30(3):804–11.

10. Young JB, Robinson M, Chell S, Sanderson D, Chaplin S, Burns E, Fear J. A whole system study of intermediate care services for older people. Age Ageing. 2005;34(6):577–83.

11. Intermediate care - half way home: updated guidance for the NHS and local authorities. Department of Health. 2009. London, England.

12. National Audit of Intermediate Care Summary Report - England. 2017. NHS Benchmarking Network. London, England.

13. Griffiths PD, Edwards MH, Forbes A, Harris RL, Ritchie G. Effectiveness of intermediate care in nursing-led in-patient units. Cochrane Database Syst Rev. 2007;2007(2):CD002214.

14. Garasen H, Windspoll R, Johnsen R. Long-term patients’ outcomes after intermediate care at a community hospital for elderly patients: 12-month follow-up of a randomized controlled trial. Scand J Public Health. 2008;36(2):197–204.

15. Garasen H, Windspoll R, Johnsen R. Intermediate care at a community hospital as an alternative to prolonged general hospital care for elderly patients: a randomised controlled trial. BMC Public Health. 2007;7:68.

16. Negm AM, Kennedy CC, Thabane L, Veroniki AA, Adachi JD, Richardson J, Cameron ID, Giangregorio A, Petropoulou M, Alsaad SM, Alzahrani J, Maaz M, Ahmed MM, Kim E, Tehfe H, Dima R, Sabanayagam K, Hewston P, Abu Alrob H, Papaioannou A. Management of Frailty: A Systematic Review and Network Meta-analysis of Randomized Controlled Trials. J Am Med Dir Assoc. 2019;20(10):1190–8.

17. Clegg AP, Barber SE, Young JB, Forster A, Iliffe SJ. Do home-based exercise interventions improve outcomes for frail older people? Findings from a systematic review. Rev Clin Gerontol. 2012;22(1):68–78.

18. Theou O, Stathokostas L, Roland KP, Jakobi JM, Patterson C, Vandervoort AA, Jones GR. The effectiveness of exercise interventions for the management of frailty: a systematic review. J Aging Res. 2011;2011:569194.

19. Prescott M, Lilley-Kelly A, Cundill B, Clarke D, Drake S, Farrin AJ, Forster A, Goodwin M, Goodwin VA, Hall AJ, Hartley S, Holland M, Hulme C, Nikolova S, Parker C, Wright P, Ziegler F, Clegg A. Home-based Extended Rehabilitation for Older people (HERO): study protocol for an individually randomised controlled multi-centre trial to determine the clinical and cost-effectiveness of a home-based exercise intervention for older people with frailty as extended rehabilitation following acute illness or injury, including embedded process evaluation. Trials. 2021;22(1):783.

20. Rockwood K, Theou O. Using the Clinical Frailty Scale in Allocating Scarce Health Care Resources. Can Geriatr J. 2020;23(3):210–5.

21. Podsiadlo D, Richardson S. The timed “Up & Go”: a test of basic functional mobility for frail elderly persons. J Am Geriatr Soc. 1991;39(2):142–8.

22. Nasreddine ZS, Phillips NA, Bedirian V, Charbonneau S, Whitehead V, Collin I, Cummings JL, Chertkow H. The Montreal Cognitive Assessment, MoCA: a brief screening tool for mild cognitive impairment. J Am Geriatr Soc. 2005;53(4):695–9.

23. Ware J, Kosinki M, Gandek B. SF36 Health Survey: Manual and Interpretation Guide. 3rd ed. Lincoln: Quality Metric Inc; 2005.

24. Collin C, Wade DT, Davies S, Horne V. The Barthel ADL Index: a reliability study. Int Disabil Stud. 1988;10(2):61–3.

25. Gladman JR, Lincoln NB, Adams SA. Use of the extended ADL scale with stroke patients. Age Ageing. 1993;22(6):419–24.

26. Herdman M, Gudex C, Lloyd A, Janssen M, Kind P, Parkin D, Bonsel G, Badia X. Development and preliminary testing of the new five-level version of EQ-5D (EQ-5D-5L). Qual Life Res. 2011;20(10):1727–36.

27. Angst F, Aeschlimann A, Stucki G. Smallest detectable and minimal clinically important differences of rehabilitation intervention with their implications for required sample sizes using WOMAC and SF-36 quality of life measurement instruments in patients with osteoarthritis of the lower extremities. Arthritis Rheum. 2001;45(4):384–91.

28. Osborne RH, Hawthorne G, Lew EA, Gray LC. Quality of life assessment in the community-dwelling elderly: validation of the Assessment of Quality of Life (AQoL) Instrument and comparison with the SF-36. J Clin Epidemiol. 2003;56(2):138–47.

29. Logan PA, Leighton MP, Walker MF, Armstrong S, Gladman JR, Sach TH, Smith S, Newell O, Avery T, Williams H, Scott J, O’Neil K, McCluskey A, Leach S, Barer D, Ritchie C, Turton A, Bisiker J, Smithard D, Baird T, Guyler P, Jackson T, Watmough I, Webster M, Ivey J. A multi-centre randomised controlled trial of rehabilitation aimed at improving outdoor mobility for people after stroke: study protocol for a randomised controlled trial. Trials. 2012;13:86.

30. Iliffe S, Kendrick D, Morris R, Griffin M, Haworth D, Carpenter H, Masud T, Skelton DA, Dinan-Young S, Bowling A, Gage H, ProAct65+ research t. Promoting physical activity in older people in general practice: ProAct65+ cluster randomised controlled trial. Br J Gen Pract. 2015;65(640):e731–8.

31. Maruish M. User’s manual for the SF-36v2 Health Survey (3rd ed.). Lincoln, RI: QualityMetric Incorporated.

32. QualityMetric, Inc., PRO CoRE 2.0 Smart Measurement System. (2020).

33. Roberts C, Roberts SA. Design and analysis of clinical trials with clustering effects due to treatment. Clin Trials. 2005;2(2):152–62.

34. Fiero MH, Hsu CH, Bell ML. A pattern-mixture model approach for handling missing continuous outcome data in longitudinal cluster randomized trials. Stat Med. 2017;36(26):4094–105.

35. Rubin D. Inference and missing data. Biometrika. 1976;63:581–92.

36. Connell AM. Employing complier average causal effect analytic methods to examine effects of randomized encouragement trials. Am J Drug Alcohol Abuse. 2009;35(4):253–9.

37. National Institute of Health & Care Excellence (NICE). NICE health technology evaluations: the manual. NICE 2022. https://www.nice.org.uk/process/pmg36/chapter/introduction-to-health-technology-evaluation Accessed 26.4.24.

38. Murray E, Treweek S, Pope C, MacFarlane A, Ballini L, Dowrick C, Finch T, Kennedy A, Mair F, O’Donnell C, Ong BN, Rapley T, Rogers A, May C. Normalisation process theory: a framework for developing, evaluating and implementing complex interventions. BMC Med. 2010;8:63.

