## Supplementary Material for "Home-based Extended Rehabilitation for Older People with Frailty (HERO): a Randomised Controlled Trial"

**Internal pilot**

**Prespecified progression criteria**

Recruitment (assessed at 6 months after the start of recruitment - June 2018).

**Green:** ≥4 patients/month/site (measured in months 4-6 to allow time for recruitment to

stabilise)

**Amber:** <4 but ≥2 patients/month/site

**Red:** <2patients/month/site

Intervention Provision (assessed at 6 months after the start of recruitment - June 2018).

**Green**: ≥80% of intervention participants receiving their first home visit within 3 weeks

**Amber**: <80% but ≥65% of intervention participants receiving their first home visit within 3

weeks

**Red:** <65% of intervention participants receiving their first home visit within 3 weeks

Intervention Acceptability (assessed at 9 months after the start of recruitment - September 2018.

**Green:** ≥80% retention of intervention participants

**Amber:** <80% but ≥65% retention of intervention participants

**Red:** <65% retention of intervention participants

Six month follow-up / completion of outcomes (assessed at 12 months after the start of recruitment - December 2018).

**Green:** ≥80% completion of the SF-36 physical component summary

**Amber:** <80% but ≥65% completion of the SF-36 physical component summary

**Red:** <65% completion of the SF-36 physical component summary

**Results and subsequent changes to trial protocol**

Recruitment and intervention provision, assessed 6 months after trial opening, were rated as green and red respectively. Overall recruitment during this period exceeded targets and no revisions to the recruitment strategy were made. Protocol amendments to improve intervention provision included extension of the recruitment window to up to 7 days post discharge, addition of researcher guidance to assist conversations with potential participants, clarification of the process for commencing intervention delivery for those participants re-admitted to hospital and the addition of further checks to ensure eligibility criteria were met prior to randomisation.

Intervention acceptability and six month follow-up, assessed 9 and 12 months after trial recruitment, were both rated as red. Protocol amendments at this stage included provision of an unconditional gift voucher at six months, addition of a participant card to prompt questionnaire completion and inclusion of a final postal contact to collect SF36 only.

Progression to the main trial was approved by the Trial Steering Committee (TSC) and Health Technology Assessment (HTA) programme.

**Outcome definitions**

**Primary outcome:**

Effect on physical health-related quality of life at 12 months.

- Mean PCS score

The SF36 consists of 36 questions used to measure 8 domains of health-related quality of life: physical functioning, social functioning, role limitations due to physical problems, role limitations due to emotional problems, bodily pain, mental health, energy/vitality, and general health perceptions. The information obtained on these eight domains can be further aggregated into two summary component measures of physical and mental health; the Physical Component Score (PCS) and Mental Component Score (MCS). The PCS score incorporates physical functioning; role-physical; bodily pain and general health scales. The scoring of the SF36 questionnaire is undertaken using OPTUM PRO CoRE software. Scores range from 0 to 100 where 0 is the worst possible health rating and 100 is the best possible health rating.

**Secondary Outcomes:**

Effect on physical health-related quality of life at 6 months.

- Mean PCS score (as defined above)

Effect on mental health-related quality of life at 6 and 12 months.

- Mean MCS score (as defined above)

Effect on activities of daily living at 6 and 12 months.

- Mean Barthel Index
- Mean NEADL Score

The Barthel index is a questionnaire designed to measure a person’s ability to care for themselves. The index covers 10 domains of self-care and aims to assess if the respondent can perform certain tasks independently. There are 2 items scored 0 or 1 (Grooming and Bathing), 6 items scored 0, 1 or 2 (Bowels, Bladder, Toilet use, Feeding, Dressing, Stairs) and the final 2 items are scored 0, 1, 2 or 3 (Transfer, Mobility). In each case a higher score indicates a greater level of independence with which the person can perform the given task. The overall score is taken as the summation of each of the individual item scores (range 0-20). Greater scores indicate greater self-care ability.

The NEADL measures help needed with instrumental activities of daily living, including walking around outside; doing the housework; using the telephone. It is a 22 item scale each with four possible responses: 0 ‘Not at all’; 1 ‘With help’; 2 ‘On your own with difficulty’; 3 ‘On your own’. All item scores are then summed to obtain the NEADL score (range 0-66). A higher score indicates greater independence.

Effect on health-related quality of life at 6 and 12 months.

- Mean EQ-5D-5L sore

EQ-5D is a 5 item, self-assessed, health related quality of life questionnaire. The scale measures quality of life including mobility, self-care, usual activities, pain/discomfort and anxiety/depression. Each item is measured on a 5-point Likert Scale, ranging between 1-no problems and 5-extreme problems. Health states are transformed into validated health utility index (range -0.594 (worst health) to 1 (best health)).

Effect on care home admission rates, hospital readmission rates, all-cause hospitalisation, hospitalisation due to falls or fractures and overall health and social care resource use at 12 months.

- Care home admission (yes/no)
- Hospital readmission (yes/no)
- Number of all-cause hospital readmissions
- Number of falls- or fracture-related hospital readmissions
- Healthcare resource use

Care home admission is the number of participants who were admitted to a care home during the trial period, obtained via a change in contact details Case Report Form.

Hospitalisation data is obtained from Hospital Episode Statistics (HES) admitted patient care (APC) dataset, accident and emergency (A&E) dataset and emergency care dataset (ECDS; replaced A&E from 2018/19) from NHS digital.

Hospital readmission is defined as the hospitalisation starting from the date of discharge home from hospital or intermediate are services as recorded on the randomisation CRF (day 0) (hospitalisation at randomisation will not be counted) or an acute inpatient admission from HES dataset (regardless of length of stay) where an acute admission will be identified through the ADMIMETH field (codes 21, 22, 23, 2A, 2B, 2D and 24).

All-cause hospitalisations will be obtained as defined above for hospital readmission.

Hospitalisations due to falls or fractures are defined as an acute inpatient admission in the APC dataset if the primary reason/diagnosis for admission included any of the ICD-10 codes W00-W19, M80, S22, S32, S42, S52, S72, S82, T08, T10, T12, T14.2 or if codes in the A&E and ECDS datasets suggest a falls-related hospitalisation.

Effect on mortality

- Deaths (yes/no)
- Time to death

Mortality data (date and cause of death) were obtained from the Civil Registrations (Deaths) Secondary Care Cut dataset and via case report form.

**Supplementary Table 1. Demographics of patients Screened, Eligible to Approach, Consented, Eligible and Randomised**

|  | **Screened**  **(n=16687)** | **Eligible to approach**  **(n=5505)** | **Consented**  **(n=905)** | **Eligible**  **(n=775)** | **Randomised**  **(n=740)** |
| --- | --- | --- | --- | --- | --- |
| **Age** |  |  |  |  |  |
| Mean (s.d.) | 83.7 (7.6) | 83.9 (7.0) | 83.0 (7.0) | 82.7 (7.1) | 82.7 (7.1) |
| Missing | 312 | 210 | 9 | 7 | 0 |
| **Gender** |  |  |  |  |  |
| Male | 6729 (40.6%) | 2000 (36.6%) | 318 (35.3%) | 269 (34.8%) | 254 (34.3%) |
| Female | 9855 (59.4%) | 3460 (63.4%) | 584 (64.7%) | 504 (65.2%) | 486 (65.7%) |
| Missing | 103 | 45 | 3 | 2 | 0 |
| **Ethnicity** |  |  |  |  |  |
| White | 15011 (92.9%) | 4779 (91.9%) | 859 (97.2%) | 739 (97.6%) | 699 (97.6%) |
| Other* | 1139 (7.1%) | 422 (8.1%) | 25 (2.8%) | 18 (2.4%) | 17 (2.4%) |
| Missing | 537 | 304 | 21 | 18 | 24 |
| **Reason for Hospital Admission** |  |  |  |  |  |
| Acute illness | 12461 (77.2%) | 3674 (70.5%) | 621 (69.5%) | 527 (68.6%) | 502 (68.5%) |
| Injury | 3688 (22.8%) | 1535 (29.5%) | 273 (30.5%) | 241 (31.4%) | 231 (31.5%) |
| Missing | 538 | 296 | 11 | 7 | 7 |

* Data have been grouped to into Other to preserve anonymity

**Supplementary Table 2. Baseline characteristics according to 12m follow-up status**

|  | Successful  (n = 479) | Unsuccessful  (n = 261) | Total  (n = 740) |
| --- | --- | --- | --- |
| Age, years | 82.5 (7.2) | 82.9 (7.0) | 82.6 (7.1) |
| Gender  Male  Female | 160 (33.4%)  319 (66.6%) | 94 (36.0%) 167 (64.0%) | 254 (34.3%) 486 (65.7%) |
| Ethnicity  White  Other* | 454 (97.4%) 12 (2.6%) | 245 (98.0%) 5 (2.0%) | 699 (97.6%) 17 (2.4%) |
| SF36 PCS Score^$^ | 31.5 (8.0) | 30.6 (8.0) | 31.2 (8.0) |
| SF36 MCS Score^$^ | 48.6 (12.0) | 48.2 (10.9) | 48.4 (11.6) |
| Barthel Index Score^$^ | 17.0 (2.6) | 16.4 (2.8) | 16.8 (2.7) |
| NEADL Score^$^ | 42.4 (14.2) | 38.5 (14.5) | 41.0 (14.4) |
| CFS Score^#^ | 5.5 (0.6) | 5.6 (0.6) | 5.5 (0.6) |
| CFS Score |  |  |  |
| Vulnerable | 1 (0.2%) | 0 (0.0%) | 1 (0.1%) |
| Mild frailty | 260 (54.4%) | 115 (44.2%) | 375 (50.8%) |
| Moderate frailty | 196 (41.0%) | 128 (49.2%) | 324 (43.9%) |
| Severe frailty | 21 (4.4%) | 17 (6.5%) | 38 (5.1%) |
| MoCA Score^$^ | 23.7 (2.9) | 23.1 (2.9) | 23.5 (2.9) |
| Reason for Admission  Acute illness  Injury | 318 (66.4%) 161 (33.6%) | 193 (73.9%) 68 (26.1%) | 511 (69.1%) 229 (30.9%) |
| Setting Discharged from  Hospital  Bed-based intermediate care  Home-based intermediate care | 166 (34.7%) 109 (22.8%) 204 (42.6%) | 92 (35.2%) 73 (28.0%) 96 (36.8%) | 258 (34.9%) 182 (24.6%) 300 (40.5%) |
| TUGT Score | 46.0 (38.9) | 47.6 (36.2) | 46.6 (37.9) |
| HOPE Level  Level 1  Level 2  Level 3 | 287 (59.9%) 128 (26.7%) 64 (13.4%) | 179 (68.6%) 59 (22.6%) 23 (8.8%) | 466 (63.0%) 187 (25.3%) 87 (11.8%) |
| Involved in previous rehabilitation programme | 31 (6.7%) | 9 (3.7%) | 40 (5.6%) |
| Number of comorbidities  None  ≥ 1 | 130 (27.4%)  344 (72.6%) | 70 (27.2%)  187 (72.8%) | 200 (27.4%)  531 (72.6%) |
| Type of comorbidity^&^  Diabetes mellitus  Chronic obstructive pulmonary disease  Congestive heart failure  Moderate to severe chronic kidney disease  Connective tissue disease  Cerebrovascular disease  Solid tumour (localised)  Solid tumour (metastatic)  Myocardial infarction  Peripheral vascular disease  Malignant Lymphoma  Dementia  Peptic ulcer disease  Leukaemia  Hemiplegia  Liver disease | 116 (34.6%)  91 (27.6%)  69 (20.9%)  73 (22.1%)  63 (19.3%)  52 (15.9%)  45 (13.6%)  2 (4.7%)  40 (12.1%)  25 (7.6%)  6 (1.8%)  6 (1.8%)  5 (1.5%)  8 (2.4%)  6 (1.8%)  2 (0.6%) | 64 (34.8%)  56 (30.8%)  50 (27.6%)  45 (24.9%)  35 (19.4%)  21 (11.6%)  31 (17.1%)  5 (17.2%)  31 (17.1%)  19 (10.1%)  6 (3.3%)  6 (3.3%)  3 (1.7%)  0 (0.0%)  0 (0.0%)  4 (2.2%) | 180 (34.7%)  147 (28.7%)  119 (23.3%)  118 (23.0%)  98 (19.3%)  73 (14.3%)  76 (14.8%)  7 (9.7%)  71 (13.9%)  44 (8.7%)  12 (2.4%)  12 (2.3%)  8 (1.6%)  8 (1.6%)  6 (1.2%)  6 (1.2%) |

Data are mean (SD) or n (%). SF36 PCS=Physical Component Score, range 0-100. SF36 MCS = SF36 Mental Component Score, range 0-100. CFS=Clinical Frailty Score, range 0-9. NEADL = Nottingham Extended Activities of Daily Living, range 0-66. Barthel Index = Barthel Index of activities of daily living, range 0-20. MoCA= Montreal Cognitive Assessment, range 0-30. TUGT=Timed Up and Go Test. *Data have been grouped to into Other to preserve anonymity. ^$^Higher scores are better. ^#^Lower scores better. ^&^Self-reported, not mutually exclusive.

**Supplementary Table 3. Eligibility violations and withdrawals**

|  | **HOPE**  **(n=410)** | **CONTROL**  **(n=330)** | **TOTAL**  **(n=740)** |
| --- | --- | --- | --- |
| **Has an eligibility violation been noted for the participant?** |  |  |  |
| Yes | 20 (4.9%) | 3 (0.9%) | 23 (3.1%) |
| No | 390 (95.1%) | 327 (99.1%) | 717 (96.9%) |
| **Has participant withdrawn from questionnaires, optional interviews or further data collection?** |  |  |  |
| Yes | 77 (18.8%) | 30 (9.1%) | 107 (14.5%) |
| No | 333 (81.2%) | 300 (90.9%) | 633 (85.5%) |

**Supplementary Figure 1. Summary of Intervention Delivery**

223 Completed Home Visit 5
 (54.4% of those randomised, 67.2% of those commencing)

243 Completed Home Visit 4
(59.3% of those randomised, 73.2% of those commencing)

283 Completed Home Visit 3
 (69.0% of those randomised, 85.2% of those commencing)

306 Completed Home Visit 2
(74.6% of those randomised, 92.2% of those commencing)

332 Commenced intervention

(81.0% of those randomised)

410 Randomised

Not commencing intervention: 78 (19.0% of randomised)

33 (42.3%) Not Appropriate

11 (14.1%) Died 
 8 (10.3%) Moved to care home
 7 (9.0%) Eligibility violators
 6 (7.7%) Unable to contact
 5 (6.4%) Referral missed
 4 (5.1%) Declined
 3 (3.8%) Withdrawn
 1 (1.3%) COVID lockdown

Initial Visit within 3 weeks: 263 (79.2% of those who commenced intervention)

Initial Visit Overdue: 69 (20.8% of randomised)

28 (41.8%) Staff Capacity
 11 (16.4%) Readmitted to Hospital
 9 (13.4%) Receiving intermediate care
 9 (13.4%) Difficulty contacting
 4 (6.0%) Holiday Period
 2 (3.0%) Weather Issues
 2 (3.0%) Change of therapist
 1 (1.5%) Eligibility violator
 1 (1.5%) Error in booking
 2 Missing

332 Completed Home Visit 1

(81.0% of those randomised, 100.0% of those commencing)

188 Completed 24 weeks of intervention delivery
(45.9% of those randomised, 56.6% of those commencing)

**Supplementary Table 3. Summary of Intervention Delivery and Content**

|  | Commenced intervention  (n = 332) |
| --- | --- |
| Timing of first home visit  Mean (SD) | 20.9 (19.02) |
| Number of contacts  Median (range) | 18 (1, 26) |
| Number of Home Visits  Median (range) | 5 (1, 9) |
| Number of Telephone Contacts  Median (range) | 14 (1, 22) |
| Level of exercise prescribed  Level 1  Level 2  Level 3  Level 1 and 2  Not prescribed exercises | 178 (53.6%) 85 (25.6%) 52 (15.7%)  1 (0.3%)  16 (4.8%) |
| Number of exercises completed per home visit  Mean (SD) | 6.0 (2.6) |
| Percentage of prescribed exercises completed  Less than 50%  Between 50% and 75%  More than 75% | 69 (21.5%) 76 (23.7%) 176 (54.8%) |
| Number who had goals set | 292 (88.0%) |
| Number of goals set  Mean (SD) | 2.2 (1.5) |
| Number of goals achieved  Median (range) | 0.0 (0, 55) |
| Time taken to achieve goals (days)  Median (range) | 35.0 (0, 283) |
| Participant engaged with weekly diary entries | 278 (83.7%) |
| Number of weekly exercise diaries completed  Median (range) | 17.5 (0, 37) |
| Length of engagement with weekly exercise diaries (weeks)  Median (range) | 22.9 (0, 129) |

**Supplementary Table 4. Summary of Usual Care services accessed**

|  | **HOPE**  **(n=410)** | **CONTROL**  **(n=330)** | **TOTAL**  **(n=740)** |
| --- | --- | --- | --- |
| **Did researcher complete a 12 month usual care review for participant?** |  |  |  |
| Yes | 204 (49.8%) | 166 (50.3%) | 370 (50.0%) |
| No | 206 (50.2%) | 164 (49.7%) | 370 (50.0%) |
| **Usual care service accessed** |  |  |  |
| Planned GP Contact | 111 (54.4%) | 89 (53.6%) | 200 (54.1%) |
| District nurse or health visitor | 108 (52.9%) | 90 (54.2%) | 198 (53.5%) |
| Practice nurse | 78 (38.2%) | 53 (31.9%) | 131 (35.4%) |
| Community matron | 52 (25.5%) | 47 (28.3%) | 99 (26.8%) |
| Physiotherapist or occupational therapist | 56 (27.5%) | 42 (25.3%) | 98 (26.5%) |
| Podiatrist/chiropodist | 43 (21.1%) | 31 (18.7%) | 74 (20.0%) |
| Other rehabilitation / therapy team member | 30 (14.7%) | 25 (15.1%) | 55 (14.9%) |
| Unplanned GP Contact | 20 (9.8%) | 19 (11.4%) | 39 (10.5%) |
| Advanced nurse practitioner | 19 (9.3%) | 15 (9.0%) | 34 (9.2%) |
| Community pharmacist | 17 (8.3%) | 6 (3.6%) | 23 (6.2%) |
| Speech and language therapist | 14 (6.9%) | 5 (3.0%) | 19 (5.1%) |
| Community mental health team | 4 (2.0%) | 6 (3.6%) | 10 (2.7%) |
| Social worker | 6 (2.9%) | 3 (1.8%) | 9 (2.4%) |

**Supplementary Table 5. Summary of Deaths and Hospitalisations due to falls or fracture**

|  | **HOPE**  **(n=410)** | **CONTROL**  **(n=330)** | **TOTAL**  **(n=740)** |
| --- | --- | --- | --- |
| **Did participant die?** |  |  |  |
| Yes | 63 (15.4%) | 62 (18.8%) | 125 (16.9%) |
| No | 347 (84.6%) | 268 (81.1%) | 615 (83.1%) |
| **Was participant hospitalised due to fall or fracture?** |  |  |  |
| Yes | 50 (12.2%) | 46 (13.9%) | 96 (13.0%) |
| No | 360 (87.8%) | 284 (86.1%) | 644 (87.0%) |
| **Number of hospitalisations due to falls and fractures** |  |  |  |
| Mean (SD) | 0.2 (0.50) | 0.2 (0.48) | 0.2 (0.49) |

**Complier Average Causal Effect (CACE analysis)**

Compliers will be defined on two levels: participant “compliance” with prescribed exercises with therapist fidelity assessed via session delivery. For the therapist delivery, the following will be defined as “compliant” in a staged approach:

1. Those who completed at least 4 home visits
2. Those who completed at least 2 home visits

Participant compliance will be assessed by the exercises completed as a proportion of the exercises prescribed:

1. Those who completed 75% of all exercises prescribed (through the entire duration of the intervention)
2. Those who completed 50% of all exercises prescribed (through the entire duration of the intervention)

The CACE analysis will take a staged approach and will be repeated four times, considering the four different levels of participant/therapist compliers, considering both therapist and participant compliance:

1. Strictest compliance:

Those who had at least 4 home visits **and** completed at least 75% of all exercises prescribed.

2a. Relaxed definition of participant compliance:

Those who had at least 4 home visits **and** completed at least 50% of all exercises prescribed.

2b. Relaxed definition of therapist compliance:

Those who had at least 2 home visits **and** completed 75% of all exercises prescribed.

3. Most lenient compliance:

Those who had at least 2 home visits **and** completed at least 50% of all exercises prescribed.

**Supplementary Table 6. CACE analysis of primary outcome, SF36 PCS score at 12 months**

|  | HOPE  (n = 410) | Effect*, 95% CI | p-value |
| --- | --- | --- | --- |
| Strictest compliance (1)  N (%) | 160 (39.0%) | -1.11 ( -3.21, 0.99) | 0.30 |
| Relaxed participant compliance (2a)  N (%) | 212 (51.7%) | -1.06 ( -3.34, 1.23) | 0.36 |
| Relaxed therapist compliance (2b)  N (%) | 173 (42.2%) | -1.35 (-3.99, 1.29) | 0.32 |
| Most lenient compliance (3)  N (%) | 240 (58.5%) | -1.31 ( -4.13, 1.52) | 0.36 |

SF36 PCS = SF36 Physical Component Score, range 0-100. *Effect represents the mean difference between treatment arms estimated using linear regression, adjusted for age, gender, previous engagement or referral to community rehabilitation services, and baseline PCS score.

**TIDieR Checklist**

Name: The Home-based Older People’s Exercise (HOPE) programme

Why (rational, theory, goal):

Frailty, loss of independence and decline in health-related quality of life become increasingly prevalent with age. These are negatively impacted by acute hospitalisation, and although periods of rehabilitation can offer some restoration of physical function and health related quality of life, these benefits may not be complete when rehabilitation ends, or sustained thereafter. Evidence indicates exercise programmes provide a positive impact physiologically, often resulting in improved mobility and function in frailty, and that behaviour change strategies positively enhance such programmes, their adoption and adherence in a population with frailty. This provides a rationale for a home-based exercise programme to extend the rehabilitation period for older people with frailty following hospitalisation with acute illness or injury. The primary goal of the HOPE programme is to improve physical health-related quality of life for older people with frailty after acute illness or injury. Secondary goals include improvements in activities of daily living, mental health, and reduced hospitalisations and care home admission.

What materials:

- The HOPE programme manual consists of five sections:

1. Information: education around exercise in older age, likely benefits and an overview of the HOPE programme
2. Safety Tips: exercise environment considerations, general precautions for exercising
3. Good Posture: guidance for maximising a good seated or standing posture for exercising
4. Exercises: level-specific exercises, each being named, information provided as to intended benefit of the exercise, bullet point text instructions for how to perform the exercise supported by photographs of a person performing the exercise.
5. Staying on Track: information provided here is aimed to help individuals remain engaged with the exercise programme long term, tips include seeking social support, making exercise fun perhaps by doing it with a friend or with music, setting goals, keeping an exercise diary, setting reminders, and what to do on a ‘bad day’.

- The manual is supplemented by a participant exercise diary, enabling the participant to monitor exercise sessions via a simple tick per session up to three times per day. The participant is also provided with a pen, to complete the diary and make notes, and a fridge magnet as a reminder to perform the exercise prescription. All materials are presented in a freestanding reusable bag, in which the items can be stored. The bag includes the HOPE programme logo as a further memory aid.
- The therapy record is provided for therapists to complete, based on a standard record used by therapy staff as part of clinical practice. It includes participant information (e.g. demographic detail, comorbidities, and medications) and individual pages for the therapy staff to record a narrative description of each participant contact (home visit and telephone calls). The therapy record pages are structured to provide some prompting to guide content/discussion at therapy sessions. The therapy record also enables recording of information required to calculate costs for intervention delivery (including travel).

What procedures:

- The HOPE programme is a 24-week home-based manualised, graded, progressive exercise intervention aimed at improving strength, endurance and balance, required for basic mobility skills like getting out of bed, standing up from a chair, walking a short distance and getting off the toilet.
- The programme is graded into three levels to account for the spectrum of frailty. The functional exercises require no special equipment, and were taught by a HOPE programme trained therapist, such that they can be performed without ongoing professional supervision.
- Participants were allocated to intervention level by:
- HOPE Level 1: Participants completing the TUGT in ≥30 seconds, who are more likely to require assistance with walking, climbing the stairs and leaving the house.
- HOPE Level 2: Participants completing the TUGT in 20-29 seconds, who demonstrate greater variability in mobility, balance and functional ability.
- HOPE Level 3: Participants who complete the TUGT in <20 seconds, who tend to be able to get in and out of a chair more easily and climb stairs.
- The programme was delivered as extended rehabilitation for trial participants allocated to receive it, upon discharge from their rehabilitation pathways following acute hospitalisation.

Who provided:

- Suitably trained and experienced community physiotherapists and therapy assistants, familiar with delivering community rehabilitation programmes to older people.
- Site physiotherapists and therapy assistants received detailed intervention training in interactive workshops delivered by trial physiotherapists experienced in HOPE programme and community rehabilitation delivery.
- Intervention training included: rationale, theory, and goals of HOPE programme, description of intervention materials and procedures, along with strategies and practical delivery of the programme.
- Supervision of therapy staff was via usual NHS line management, with intervention delivery support/advice provided by the central trial team.
- Access to training materials was available to trained therapists, and regular updates and communication with trained therapists was maintained by the central trial teams.
- Communication between trial therapists for sharing of experiences and learning was facilitated by the central trial team, through regular therapist teleconferences, site update meetings and newsletters.
- Therapists delivering the HOPE programme did not treat usual care participants referred to community rehabilitation during the study period where feasible.

How and where: mechanism and location of delivery

- The HOPE programme was delivered by the trained therapists via one to one home visits and telephone contacts for ongoing support.
- Individual home visits and telephone calls were scheduled weekly, but in a flexible manner to fit participant availability.
- No specialist rehabilitation equipment was required.

When and how much:

- The HOPE programme was delivered over a 24 week schedule including five face-to-face home visits, and nineteen telephone contacts for ongoing support.
- The exercise routines typically take less than 15 minutes to complete, and participants were requested to complete the routine three times per day on five days of the week (as able).
- A graded approach was taken to build up the exercise prescription in line with participants physical capacity.
- Progression of the programme included increased repetition of a given exercise prescription, introduction of additional exercises from the HOPE manual, or progression to the next level within the programme.
- Review of exercise performance occurred at each weekly contact, and appropriate adjustment of the exercise prescription made.
- Participants documented exercise session completion in the exercise diary.

Tailoring:

- The initial HOPE programme level was tailored to an individual in line with TUGT at baseline.
- Physiotherapists further tailored the programme on an ongoing weekly basis in line with the physical capacity and health status of the individual participant.

Modifications:

- The HOPE programme was a 24 week programme, which included an additional twelve weeks of telephone support when compared to the originally piloted HOPE programme.
- The trial was impacted by a period of national lockdown due to the COVID-19 pandemic. During this time (March 2020-Oct 2020) the trial paused to recruitment and face-to-face intervention delivery. Some participants in receipt of the HOPE programme were impacted:
  - Those recruited/randomised and who had not received ≥2 home visits to teach and check the intervention exercise performance, were discontinued from the intervention (n=8).
  - Those who had started the intervention, having received ≥2 but <5 home visits, had their remaining home visits switched to telephone contacts (n=14). This limited the progression of exercise such that no new exercises were taught and only the dose of the previously taught exercises could be adjusted.

How well (planned):

- Exercise diary and therapy record captured data regarding the exercise prescription, exercise performance and exercise session completion.
- Therapy contact data were recorded including the personnel performing, date and duration, mode of delivery, content of session.
- Process evaluation activity included review of intervention fidelity. This activity included non-participant observations and review of the training sessions completed, non-participant observations of implementation of the HOPE programme at home visits and telephone contacts, and participant, carer, therapist and therapy service manager interviews across a purposeful sample.

How well (actual):

- Of the 410 trial participants randomised to receive the intervention, 332 commenced the HOPE programme. The main reasons for not initiating the programme were:
  - Therapist deemed no longer appropriate (42.3%)
  - Participant had died (14.1%)
- Other key intervention delivery metrics are available in supplementary table 3 and supplementary figure 1.

The process evaluation concluded that the intervention had been delivered broadly as planned without significant variation between sites/regions.
